# Data Auditing and Quality Assurance in a Federated Learning Consortium; Getting the Best of Both Worlds from Cross-Institutional and In-House Data Quality Inspection

**DOI:** 10.64898/2026.09.08.26361592

**Authors:** J. (Joshi) Hogenboom, N. (Natacha) Perez, Q. (Quentin) Filori, A. (Alric) Sans, A. (Aiara) Lobo Gomes, A.L.A.J. (Andre) Dekker, W.T.A. (Winette) Van Der Graaf, O. (Olga) Husson, H. (Hugo) Crochet, L.Y.L. (Leonard) Wee, V. (Varsha) Gouthamchand

**Author notes:** Authors contributed equally and share first authorship (J. (Joshi) Hogenboom, N. (Natacha) Perez).

## Abstract

**Introduction:** Rare and heterogeneous disease research increasingly relies on privacy-enhancing technologies such as federated learning (FL) to enable cross-institutional collaboration across fragmented datasets. However, data quality assurance in FL can be limited, as individual-level data may not be directly accessible. While schema-dependent inspection offers a partial solution, it requires standardised schemas or resource-intensive frameworks, hindering scalability and collaboration. To address these challenges, we implemented two complementary dashboards and evaluated their interplay.

**Methodology:** We adapted a recognised data quality control framework for re-use of electronic health record (EHR) data using breast cancer records from Centre Léon Bérard into two dashboards: (1) an in-house dashboard for intra-clinic quality assessment, and (2) a federated dashboard for inter-clinic quality assessment. Artificial inconsistencies were introduced into distributed datasets mirroring the inhouse source to evaluate detection capabilities.

**Results:** The in-house dashboard provided granularity and reliability, pinpointing individual-level inconsistencies, while the federated dashboard enabled cross-institutional pattern detection – trade-offs inherent to their designs. The federated system revealed ecosystem-wide trends inaccessible to single-institution tools, whereas the in-house dashboard provided local validation and thoroughness.

**Discussion:** Our findings confirm a complementary relationship: FL dashboards provide scalable, collaborative oversight but may require additional quality checks, while in-house tools ensure thoroughness at the cost of scalability. A combined model seemingly offers the optimal balance, accommodating both institution-specific needs and collaborative research requirements in evolving, multi-institutional ecosystems.

## Introduction

Rare and atypical healthcare demographics expose a fundamental weakness in the current architecture of clinical research, requiring adapted strategies as traditional approaches often fail to deliver timely, generalisable evidence. Their rarity reflects not only low prevalence, but also fragmented expertise and a persistent absence of robust comparative data. To obtain meaningful insights into such heterogeneous populations, researchers typically require large-scale, geographically dispersed datasets to aggregate a large number of cases. However, such collaboration is complicated by data protection legislations and the ethical imperative to safeguard individual privacy. This tension between the need for comprehensive, multi-regional data access and the non-negotiable protection of the right of individual privacy has motivated the development of new analytical processes. Thereto, Federated Learning (FL) is a recently developed privacy-enhancing technology (PET) (1) that can facilitate such geographically dispersed and privacy-aware ambitions.

Generally, FL enables analyses across distributed data cohorts by aggregating local computations into a global result. In its naivest illustration, a “federated average” is trivially easy to compute for two equal-sized horizontally partitioned datasets with local averages of 20 and 30 respectively; that is, the global average is simply the arithmetic average of the local averages, thus the global answer is 25. In general, what a federated statistic calculates will be the same as if (hypothetically) all of the distributed data will be gathered in a single repository. Naturally, due to privacy, this might not be achievable in every possible situation.

As a PET – though implementation-dependent – FL shields individual patient-level data, addressing privacy, and regulatory constraints, but allows mathematically correct calculations to be done without physically transferring patient data. By lowering some of the legal and organisational barriers about data access, FL has potential to accelerate multi-centre research, whereas centralised approaches must apply more caution due to potential re-identification risks.

Conversely, FL’s privacy-by-design measures might have important consequences decelerating data quality assurance. Without direct access to individual-level data, identifying outliers, filtering implausible values, and other data quality issues can become significantly more challenging to detect (2). Consequently, what FL gains in terms of collaboration and more diverse population size, it also sacrifices in highly granular quality inspection capability.

To address this limitation, robust data quality frameworks are essential. Data quality often refers to the degree to which data meets context-dependant requirements on dimensions as completeness, conformance, and plausibility, with intra-clinic and inter-clinic contexts requiring distinct approaches. However, their adoption is hindered by heterogeneous data models between institutions, as tools can rarely be applied across different schemas. The global data model landscape remains a shattered mosaic despite harmonisation efforts (e.g. HL7 FHIR (3), OpenEHR (4), OSIRIS (5), OMOP (6) and data quality initiatives like QUANTUM (7)). Consequently, most tools remain confined to isolated, time-constrained silos due to the need for stable underlying data models and thus are ill-equipped for multi-party collaboration.

These challenges are particularly acute in the context of electronic health records (EHR) data, which, while un-curated and ill-suited for research without cleaning, reflect real-world complexity and potential. To harness this potential, dedicated quality assessment frameworks, such as that by Kahn et al. (8) define completeness, conformance, and plausibility as key dimensions for intra-clinic evaluation of data integrity within individual institutions using raw, local data. However, as Declerck et al. (9) highlight, such frameworks are often re-interpreted and contextually adapted. For FL, they must also be adapted: intra-clinic dimensions such as Kahn’s typically rely on individual-level value assessments, while inter-clinic quality operates on aggregates, ought to evaluate metrics between institutions and assess attribute harmonisation through consistency and semantic alignment. Both intra- and inter-clinic levels appear essential for comprehensive quality assessment to avoid bias from heterogeneous practices or coding systems.

These challenges – balancing standardisation with contextual adaptability – are especially pronounced when implementing quality assessment tools in a genuine FL infrastructure like STRONG AYA. The STRONG AYA initiative is a living, evolving FL infrastructure for Adolescent and Young Adult (AYA) cancer, integrating historical registry data, EHRs from participating institutions, and prospectively collected longitudinal research data. All data collated for STRONG AYA adheres to a consensual set of elements defined by the AYA Core Outcome Set (COS) (10), which specifies the variables to be included without prescribing a rigid data model. Given the diversity of data sources, STRONG AYA ensures its data is F.A.I.R. (Findable, Accessible, Interoperable, Reusable) (11) and adopts semantic interoperability rather than enforcing a single model (12). Aligned with the Personal Health Train manifesto (13), STRONG AYA’s FL infrastructure prohibits individual-level data from leaving local premises, inherently preventing direct inspection and necessitating exploratory, aggregate-level quality analyses. The combination of heterogeneous data sources, varying quality, formats, and federated privacy constraints renders traditional quality control approaches inadequate. Thus, a federated quality assessment approach is essential to enable consistent, privacy-aware insights across this evolving multi-institutional infrastructure.

This work demonstrates a working implementation of different levels of data quality control possible within this complex federated consortium framework. We developed and compared two exploratory data quality inspection dashboards: one designed for in-house (intra-clinic) analysis, and another intended for federated (inter-clinic) settings. We test the hypothesis that both intra- and inter-clinic levels are essential and complementary for comprehensive quality assessment in consortia such as STRONG AYA, with the in-house dashboard best suited to detect individual-level errors (e.g. plausibility) due to its access to raw data, and the federated dashboard best suited to identify cross-institutional patterns (e.g. systematic biases) due to its reliance on aggregate outputs. Our comparison concludes with an interest holder-driven STRONG AYA use case, investigating how these approaches can tackle the FL challenges while supporting reliable multi-institutional research.

## Methodology

### Data description and processing

The data used in this study was extracted from the EHR at Centre Léon Bérard (CLB), a comprehensive cancer centre in Lyon, France, dedicated exclusively to oncology care and research. The study used a subset of breast cancer patients recorded in the institutional breast cancer database, restricted to patients with an information level of at least 5 – a threshold corresponding to explicit patient consent for the use of their health data in research. Secondary use of these healthcare data was conducted in accordance with the French MR004 reference methodology. The project underwent review by the data protection office of CLB, received a General Data Protection Regulation (GDPR) certificate, and was registered in CLB’s GDPR registry. To limit re-identification risks inherent to routine clinical data reuse, we applied an internal privacy transformation involving uniform date-shifting, thereby maintaining internal temporal relationships between events. All data processing procedures are documented in the information notice available on the Unicancer transparency portal (14), for review by individuals included in the dataset.

We selected a cohort of pathologically confirmed breast cancer patients aged 18-39 at diagnosis who received surgical treatment at CLB between 29 February 1996 and 15 February 2024. This dataset was processed through CLB’s automated extract-transform-load (ETL) pipeline, which converted the native EHR data model into the OSIRIS Real World Data model OSIRIS-RWD (15) is an in-development adaptation of the standard OSIRIS model optimised for real-world data integration and clinical applications.

To ensure that we could evaluate the detection capabilities of both the in-house and federated dashboards, we introduced controlled inconsistencies representing two categories of quality issues: intra-clinic errors to test the in-house dashboard’s ability to detect individual-level inconsistencies, including temporal discrepancies in age at diagnosis and last known vital status, as well as semantic conflicts involving implausible combinations of cancer topography, biological sex, laterality, and

TNM-UICC classification; and inter-clinic errors to test the federated dashboard’s ability to detect cross-institutional patterns, including biases in coding practices, such as using two different neoplasm topography coding systems across sites, as detailed in *Supplementary Table 1*.

To simulate a real-world scenario for our in-house and federated dashboards, we created two distinct copies of this pre-processed dataset. The first copy preserved the entire cohort of 839 patients and was imported into the in-house Microsoft SQL server. For the federated setting, we generated two variants from the second copy to simulate data from two independent centres: one maintaining the original sample size and another randomly reduced to 790 patients. Date information was transformed into relative time differences across both federated variants, and specific elements were relabelled in one variant (e.g. converting ICD-10 tumour topography codes to ICD-O-3) to introduce controlled schema heterogeneities between federated data stations. These modifications reflect real-world federated settings, where data-sharing agreements often require date obfuscation and where participating centres frequently adhere to heterogeneous data schemas.

Whilst both copies adhered to the OSIRIS-RWD standard, FL systems routinely deal with diverse data models and concept terminologies (2). To reflect common FL practice, we F.A.I.R.-ified both datasets using the Flyover tool (16), which converts structured data into RDF triples (17), annotates them with standardised terminologies for semantic interoperability (2, 12), and stores the resulting graphs in a graph database for SPARQL querying (18). To complete the FL simulation, we deployed a local Vantage6 (19, 20) developer network with a central server connecting to two data stations, each hosting one of the aforementioned graph database instances.

### Data quality framework adaptation

Our data quality assessment framework was structured around three core dimensions adapted from Kahn et al. (8): ‘Completeness’, ‘Consistency’, and ‘Semantic Consistency’. To accommodate the distinct requirements of *intra-clinic and inter-clinic quality assessment*, we tailored these dimensions accordingly. For intra-clinic assessment, we applied Kahn’s definition of ‘Semantic Consistency’ to validate logical coherence within the dataset. Whereas for inter-clinic assessment, we repurposed this dimension to assess semantic interoperability – critical for evolving, heterogeneous ecosystems such as STRONG AYA – ensuring we could evaluate what data was made semantically interoperable across independent data sources. The semantic consistency validation rules, grounded in the European Commission’s Joint Research Centre guidelines (21) were primarily designed for the intra-clinic assessment to validate data coherence, and to lesser degree were utilised for inter-clinic quality assessment. These guidelines define clinically implausible combinations of demographic and tumour characteristics (e.g. age-morphology-topography and sex-topography). The ENCR-recommended validation rules, distributed as CSV files, were directly integrated into our quality assessment tools and are designed for regular updates to reflect periodic revisions to the published recommendations (22).

### Dashboard development

For the in-house implementation, we developed a dashboard using Microsoft Power BI, leveraging its DAX language capabilities to perform aggregations and relational joins on the Microsoft SQL database. The dashboard incorporated all quality control metrics and interactive visualisations, with Power BI Report Server facilitating web-based deployment within CLB’s internal infrastructure. This implementation enabled direct querying of the pre-processed dataset while maintaining full access to individual-level data for comprehensive quality assessment.

The federated dashboard was constructed using Plotly Dash and integrated with the Vantage6 software through Vantage6’s Python client. Unlike typical federated analyses that aggregate results, our implementation was designed to retrieve and display data station-specific information. Upon initialisation, the Dash application launches two distinct federated tasks: one to identify available data concepts in each data station, and another to collect general statistics including missing and implausible value counts for key concepts. Key concepts were extracted from the JSON-LD mapping, with inlier detection manually defined for this study; in future, such constraints could also be fetched from JSON-LD when it supports them. This approach ensures individual data station visibility while operating in the privacy-aware space; a schematic of the FL-task integration is available in Supplementary Figure 1. The underlying federated algorithms are open-source and available on GitHub (23).

The complete methodological workflow, including dataset separation, processing pipelines for both in-house and federated approaches, and their respective data handling procedures, is summarised in Figure 1.

**Figure 1:**
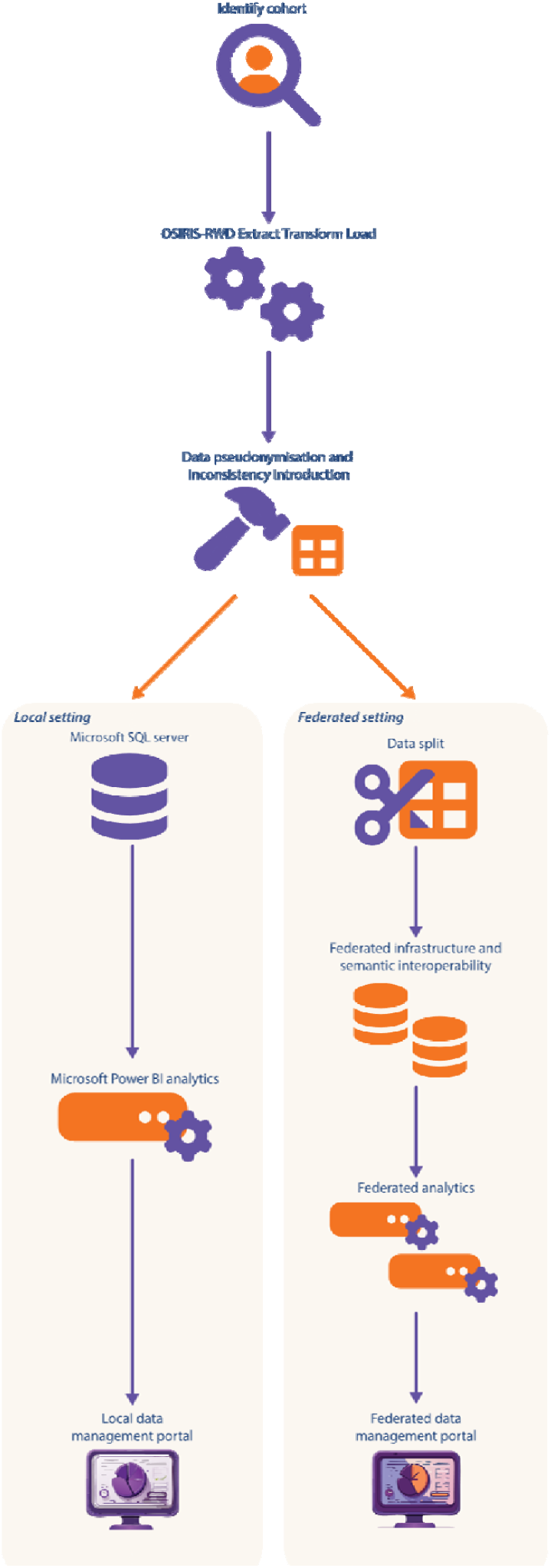
The complete workflow of the deployment of the in-house and federated dashboard for this evaluation.

### Demonstration of usability

To demonstrate the practical utility of the developed dashboards, we conducted a manual feasibility assessment of two interest holder-driven use cases (24) from the STRONG-AYA consortium. Using the implemented data quality framework, we evaluated whether the available data elements in the ingested data would support preliminary analyses of: 1) 1-, 3-, and 5-year survival rates among AYA cancer patients, and 2) the proportion of patients receiving high-cost cancer treatments. This assessment focussed on determining whether the dashboards could provide sufficient high-level insights into data completeness, consistency, and semantic validity to inform the potential executability of these analyses, rather than performing the actual analyses themselves.

## Results

Both data quality dashboards provided general information such as the total number of available patients in the dataset(s). The in-house dashboard displays the last quality control update timestamp, the number of unique patients with inconsistencies, and the absolute count of patients with complete oncology data records, as shown Figure 2. The federated dashboard included extensive information on data origin – per organisation and country – but did not display explicit data update dates or overall dataset completeness metrics, instead showing complete datapoints in donut charts as in Figure 3.

**Figure 2:**
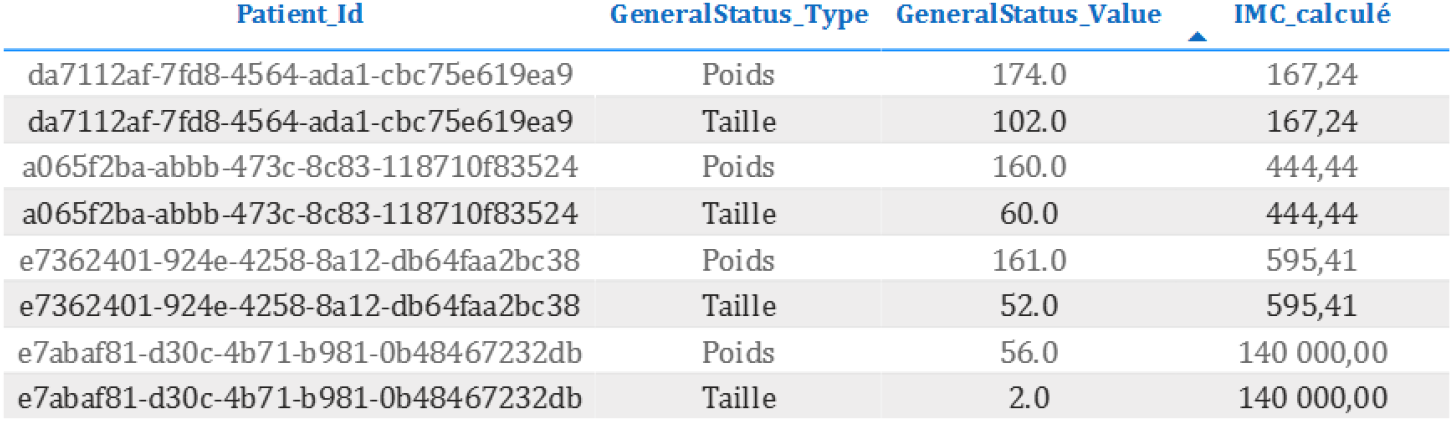
Dataset-level quality indicators and atemporal inconsistencies in the in-house dashboard (inconsistent weight and height values). ‘Poids’ represents weight in kilograms, ‘Taille’ represents length in centimetres, and ‘IMC_calculé’ represents calculated Body Mass Index. All values use ‘,’ (comma) as decimal sign.

**Figure 3:**
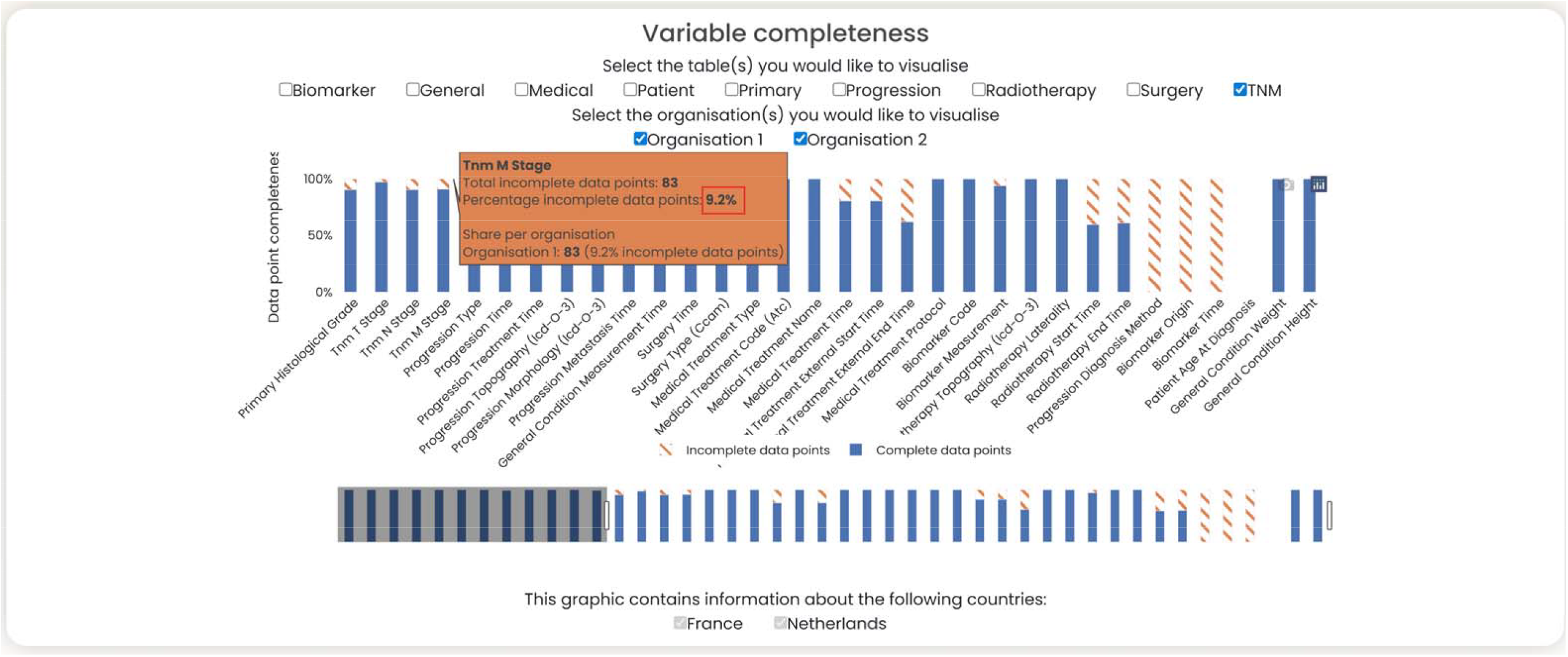
Data Completeness rate in federated dashboard: proportion of missing UICC TNM M values (highlighted in the red box).

## Metric-by-metric comparison

Completeness was generally assessed in a similar manner in both dashboards and revealed comparable results, which we exemplified through the Union for International Cancer Control’s Tumour-node-metastasis classification (UICC TNM) metastasis (M) value completeness rate with 9.2% incomplete data points, illustrated in Figure 3 (marked by the red box). Both dashboards used bar charts for this evaluation but implemented them in different ways: the in-house dashboard showcased the absolute quantity of missing data, whereas the federated dashboard explicitly displayed it as a proportion of the total number of data points. The in-house dashboard enabled granular assessments of completeness, providing insights at the individual and variable levels, including oncology-critical fields (e.g. primary cancer topography code). The federated dashboard, limited to aggregate data, focussed on inter-organisational insights through variable-by-variable bar charts, enabling assessment of whether missingness was variable-specific or institution-specific.

Atemporal consistency assessments were possible in both dashboards. For example, BMI derived from weight and height measurements revealed 4 implausible values (0.6% of entries), highlighted in Figure 2 were successfully identified in both dashboards. Notably, inconsistent values were traceable on an individual level using the corresponding patient identifier with the in-house dashboard, whereas such anomalies were not available in the federated context given the restrictive privacy conditions.

For temporal consistency, the in-house dashboard included dedicated verification of the chronological coherence between events (e.g. cancer diagnosis dates and treatment initiation dates). The federated dashboard did not have a dedicated section for this but relied on time differences, where negative values signified temporal inconsistencies.

Semantic consistency sections demonstrated distinct applications in each dashboard. The in-house dashboard identified implausible data combinations, such as 4 female patients diagnosed with epididymal cancer (ICD-10 code C63.0, a male-exclusive site per ENCR guidelines) as illustrated in Supplementary Figure 2 (available on request, in French). Due to privacy constraints, the federated dashboard could not perform individual-level assessments but instead focussed on semantic interoperability, evaluating the availability and usability of data across organisations. This was exemplified in the semantic inconsistency of neoplasm topography coding systems highlighted in Figure 4 (marked in the red box) – indicating how organisation 1 uses ICD-O-3, and organisation 2 uses ICD-10.

**Figure 4:**
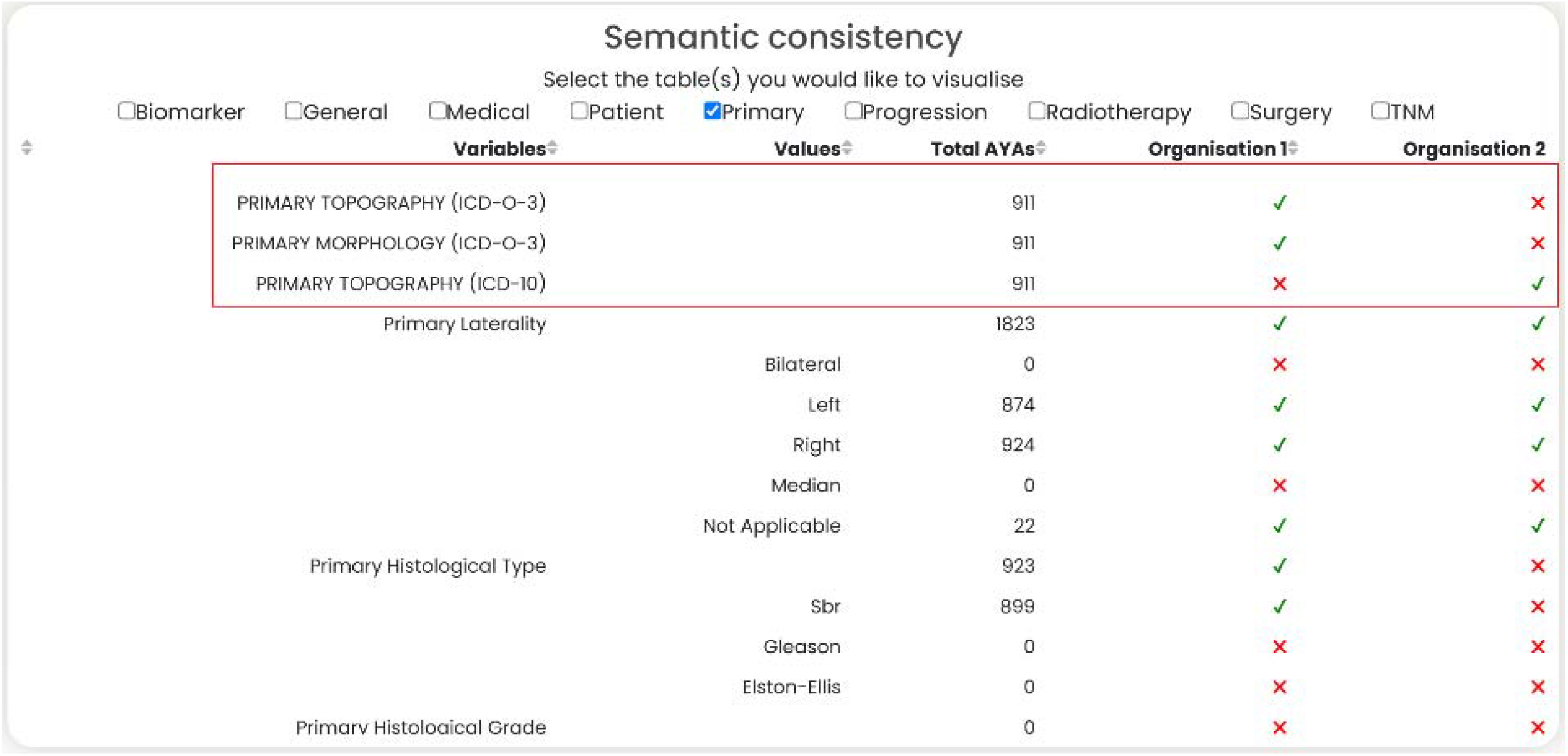
Inter-organisational data availability and semantic interoperability assessment within the federated dashboard: different neoplasm topography coding systems (highlighted in the red box).

The federated dashboard also enabled detection of non-independent and identically distributed (non-i.i.d.) value distributions between centres. For example, as illustrated in Figure 5, the dashboard identified inter-clinic heterogeneity in the availability of laterality recordings, highlighting a systematic difference in populations – albeit artificial – across participating sites. This capability addresses a key challenge in federated settings, where undetected inter-clinic variations can introduce bias and calculation errors. The full displays of the respective dashboards are available in Supplementary figures 3 to 5 (Supplementary figure 3 is available on request, in French).

**Figure 5:**
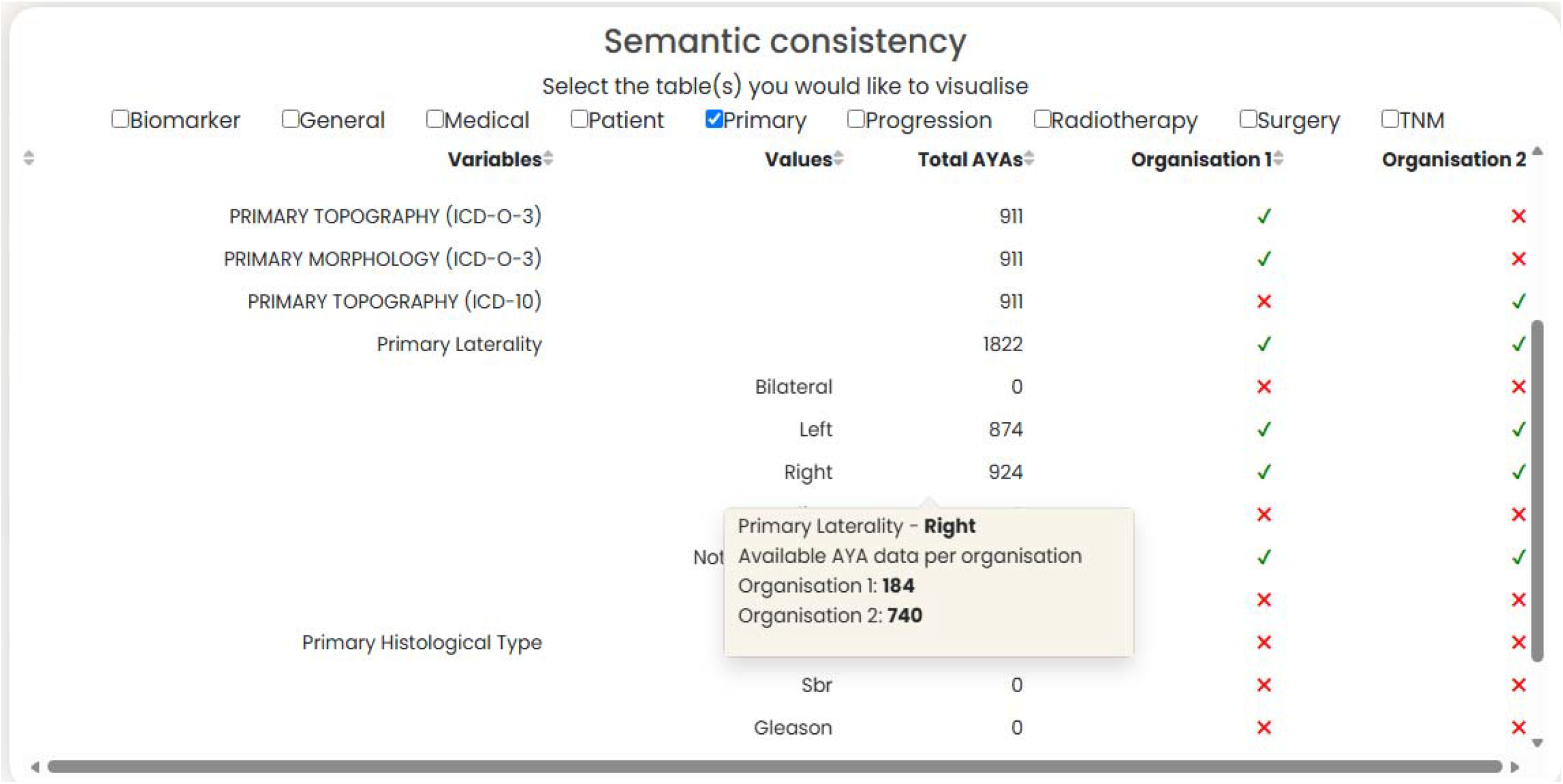
Semantic consistency assessment illustrating organisational heterogeneity through the difference in recordings of Primary laterality; in which organisation 1 has predominately left-sided cancers (not explicitly shown) and organisation 2 has predominantly right-sided cancers.

We summarised the differences for our in-house and federated dashboards in table 1, demonstrating the complementary strengths of the dashboards: the in-house dashboard’s ability to trace errors to individual patients, and the federated dashboard’s capacity to provide cross-organisational insights.

**Table 1:**
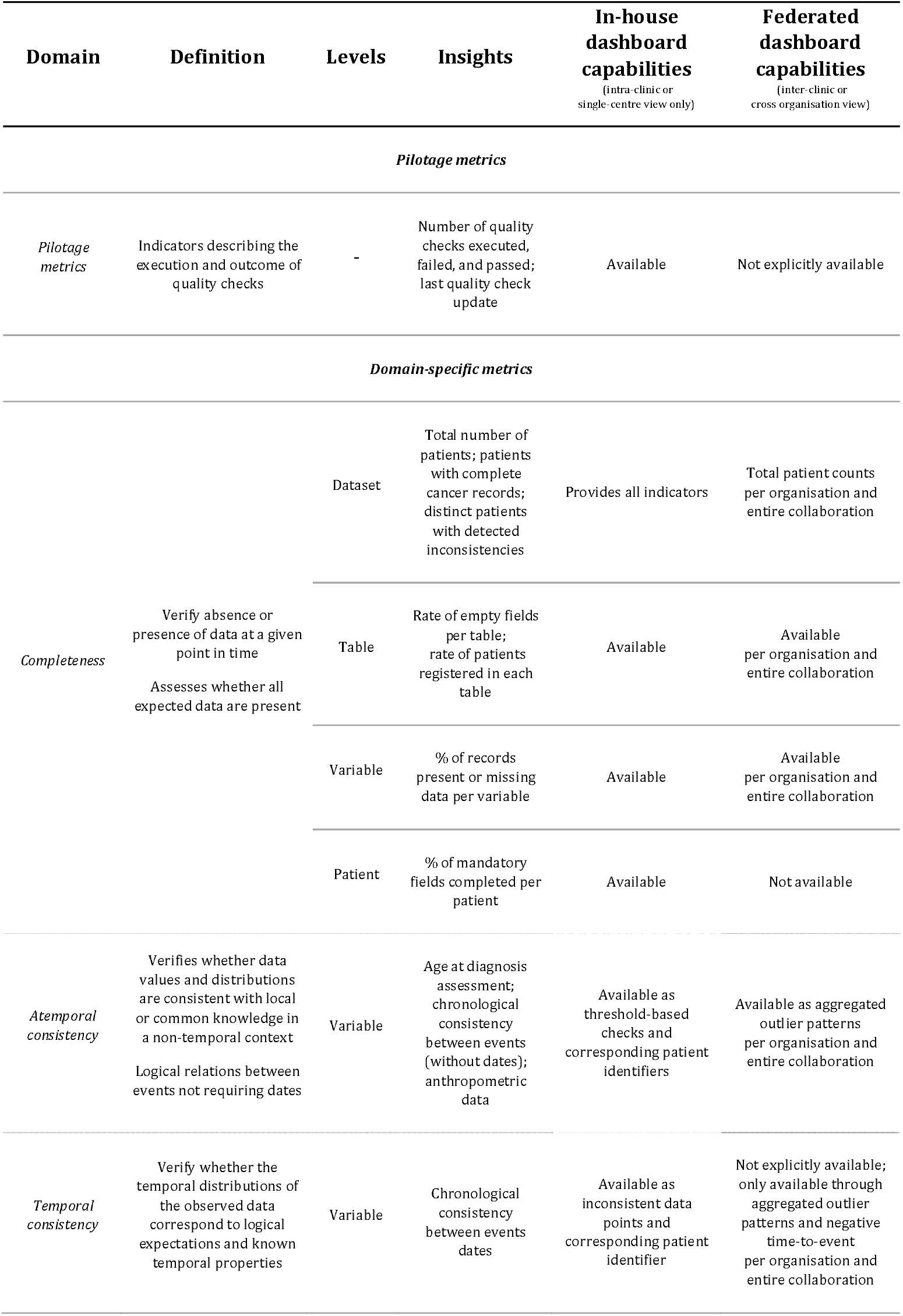
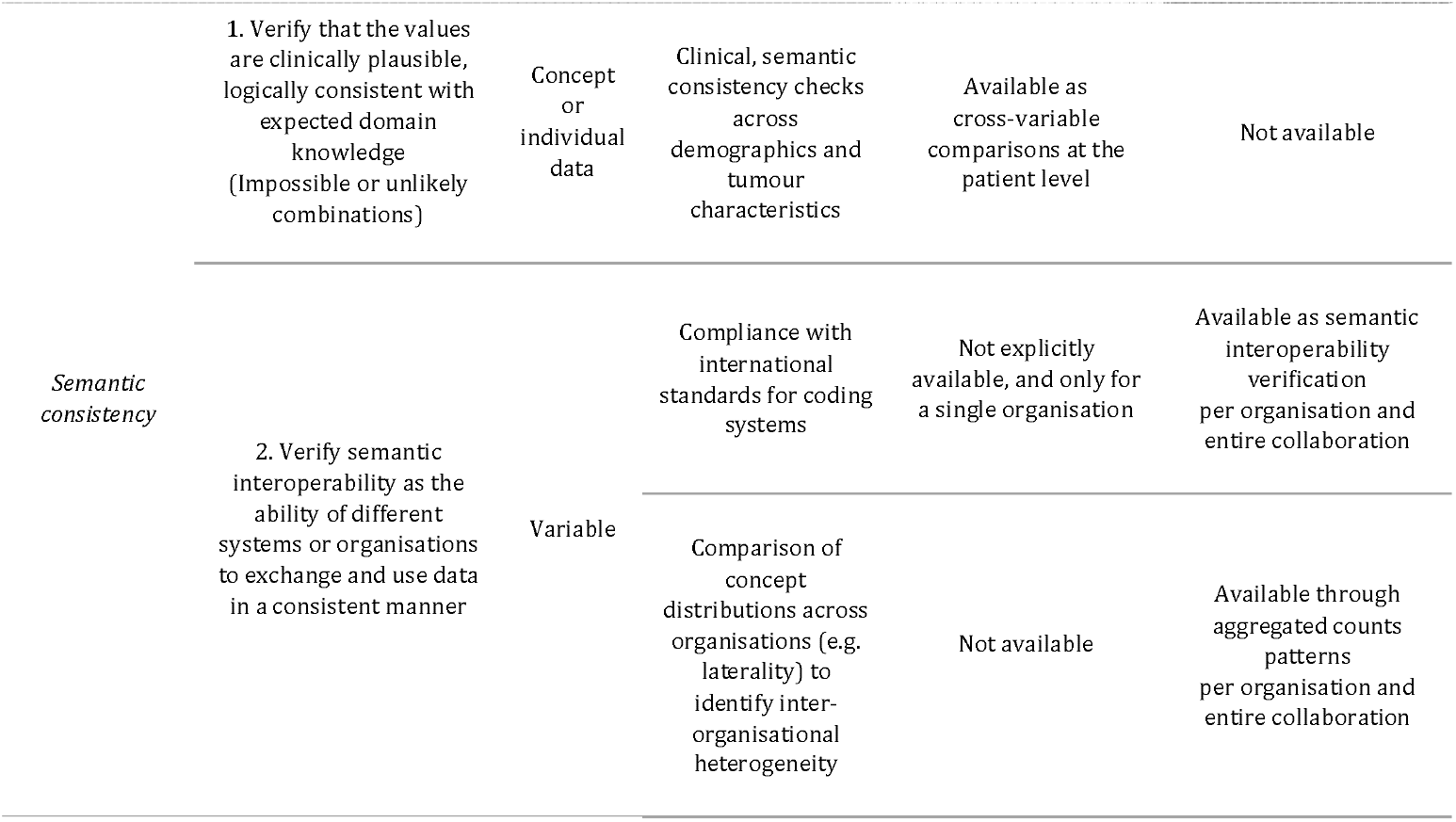
Differential Data Quality assessment capabilities of in-house versus federated dashboards.

| Domain | Definition | Levels | Insights | In-house dashboard capabilities<br>(intra-clinic or single-centre view only) | Federated dashboard capabilities<br>(inter-clinic or cross organisation view) |
| --- | --- | --- | --- | --- | --- |
| *Pilotage metrics* |  |  |  |  |  |
| *Pilotage metrics* | Indicators describing the execution and outcome of quality checks | - | Number of quality checks executed, failed, and passed; last quality check update | Available | Not explicitly available |
| *Domain-specific metrics* |  |  |  |  |  |
| *Completeness* | Verify absence or presence of data at a given point in time<br>Assesses whether all expected data are present | Dataset | Total number of patients; patients with complete cancer records; distinct patients with detected inconsistencies | Provides all indicators | Total patient counts per organisation and entire collaboration |
|  |  | Table | Rate of empty fields per table; rate of patients registered in each table | Available | Available per organisation and entire collaboration |
|  |  | Variable | % of records present or missing data per variable | Available | Available per organisation and entire collaboration |
|  |  | Patient | % of mandatory fields completed per patient | Available | Not available |
| *Atemporal consistency* | Verifies whether data values and distributions are consistent with local or common knowledge in a non-temporal context<br>Logical relations between events not requiring dates | Variable | Age at diagnosis assessment; chronological consistency between events (without dates); anthropometric data | Available as threshold-based checks and corresponding patient identifiers | Available as aggregated outlier patterns per organisation and entire collaboration |
| *Temporal consistency* | Verify whether the temporal distributions of the observed data correspond to logical expectations and known temporal properties | Variable | Chronological consistency between events dates | Available as inconsistent data points and corresponding patient identifier | Not explicitly available; only available through aggregated outlier patterns and negative time-to-event per organisation and entire collaboration |
| Semantic consistency | 1. Verify that the values are clinically plausible, logically consistent with expected domain knowledge (Impossible or unlikely combinations) | Concept or individual data | Clinical, semantic consistency checks across demographics and tumour characteristics | Available as cross-variable comparisons at the patient level | Not available |
|  | 2. Verify semantic interoperability as the ability of different systems or organisations to exchange and use data in a consistent manner | Variable | Compliance with international standards for coding systems | Not explicitly available, and only for a single organisation | Available as semantic interoperability verification per organisation and entire collaboration |
|  |  |  | Comparison of concept distributions across organisations (e.g. laterality) to identify inter-organisational heterogeneity | Not available | Available through aggregated counts patterns per organisation and entire collaboration |

### Demonstration of usability

Both dashboards enabled assessment of data feasibility for the interest holder-driven use cases by evaluating availability, completeness, and plausibility.

For the survival analysis use case, both dashboards allowed verification of survival status completeness and the availability or calculability of survival time through consistency assessments. The federated dashboard had limitations in providing distinct temporal consistency information, and traceability of erroneous data was limited compared to the in-house dashboard. For example, the in-house dashboard could identify exactly which event times were flawed, whereas the federated dashboard could only indicate that a given number were problematic. Conversely, the federated dashboard enabled aggregation of time-to-event data across institutions without disclosing individual-level information, while the in-house dashboard was limited to single-centre estimates.

For the high-cost treatment use case, both dashboards helped verify whether treatment information was available and whether its recording was complete, thereby increasing confidence in the feasibility of the analysis. Although financial cost data are typically not accessible or standardised, the federated dashboard provided an overview of treatment distribution patterns across institutions, highlighting inter-site variations in the use of intensive therapies. The in-house dashboard complemented this with greater granularity, such as linking high-cost treatments to patient-level variables, comorbidities, or outcomes. It also offered a unique advantage in scenarios requiring identification of patients eligible for in-depth longitudinal analysis by evaluating patient-level completeness and temporal plausibility across clinical events – a capability not feasible with the federated dashboard due to confidentiality constraints, which instead ensured privacy by avoiding individual-level access.

## Discussion

In this work, we evaluated the effectiveness of established data quality assurance guidelines in both intra-clinic in-house and inter-clinic federated contexts within the STRONG-AYA infrastructure.

In comparing intra-clinic in-house and inter-clinic federated dashboards for data quality assessment, we found both approaches viable, each with distinct strengths and weaknesses. While the intra-clinic in-house dashboard offers higher granularity and traceability, the inter-clinic federated dashboard is not allowed to display such granularity on potentially-flawed data points due to privacy constraints. From an intra-clinic perspective, this is a limitation. However, from an inter-clinic perspective, solely using in-house dashboards lack scalability and generalisability. Inter-clinic federated dashboards uniquely enable cross-institution quality assessment, such as detecting non-independent and identically distributed data — a critical aspect in FL — while serving all partners and highlighting flaws at scale. Conversely, intra-clinic in-house dashboards allow institutions to incisively rectify erroneous data points. Together, they form a complementary relationship, with each addressing distinct yet critical quality dimensions. To situate these findings, we consider the broader challenges of standardisation in FL ecosystems.

In the broader context of FL quality assessment, standardisation presents challenges. While uniform dashboards could theoretically standardise quality assessment, their implementation is hindered by persistent interoperability challenges. The lack of semantic and syntactic standardisation across parties makes uniform deployment impractical without substantial resource investment.

Adhering to data models such as OSIRIS-RWD or OMOP-CDM reduces these challenges, enabling in-depth quality evaluation solutions like the in-house dashboard developed here or OMOP’s ‘Data Quality Dashboard’ (25) and ‘Achilles’ (26). For example, in the OncoDataShare initiative (27), data structured using OSIRIS-RWD (with ongoing efforts to align with OMOP-CDM) enables both local and collective quality monitoring, demonstrating that standardised data models enable cross-institution insights, not just dashboard re-use.

However, enforcing such schemas places a substantial burden on non-adherent contributors and introduces rigidity. Mapping heterogeneous local systems to a shared model can introduce approximation, information loss, or and similar mapping errors. In contrast, the FL dashboard avoids these risks, revealing inter-clinic heterogeneities in terminologies and standards.

Acknowledging that FL inherently requires addressing data interoperability, semantically interoperable graph databases (2, 12) provide a natural solution to overcome these hurdles, enabling uniform evaluation solutions such as SHACL (28) validation. However, SHACL’s rigidity, while enforcing precise data rules, makes it unsuitable for technically valid but unusual data points. In contrast, a dashboard approach allows any data to be ingested, including erroneous data, but requires re-ingestion after rectifications. This flexibility resembles STRONG AYA’s pragmatic approach, which prioritises broad data inclusion over strict validation. Regardless, while intra-clinic dashboards could address some of these data quality limitations through higher granularity, they would hinder inter-clinic insights and reduce collaborative opportunities.

### Strengths and limitations

Despite these challenges, shared adherence to Kahn et al.’s guidelines across intra-clinic in-house and inter-clinic federated dashboards offers unique insights into their complementary strengths. While in-house dashboards provide comprehensive, highly granular quality assessment within individual institutions, the FL dashboard’s schema-agnostic design enables scalable deployment across diverse collaborations. This federated approach, despite privacy constraints, reveals collaboration-wide patterns – such as non-IID distributions – that remain invisible to intra-clinic tools. The trade-off suggests a future direction: combining the FL dashboard’s schema-agnostic approach with in-house dashboards’ detailed inspection to create a robust, schema-agnostic quality assessment combination. Here, the FL dashboard facilitates inter-institutional comparisons, complementing in-house assessments without matching their depth.

The metadata-dependent approach grants the FL dashboard flexibility across ecosystems beyond STRONG AYA’s implementation. We present working versions, though data requirements across projects may necessitate emphasizing different aspects, such as non-IID visualisation. This specificity extends to both inter-clinic and intra-clinic settings, where design choices must align with each context’s unique demands – as Declerck et al. already acknowledged (9). Moreover, privacy requirements in FL remain context-specific, with interpretations varying across collaborations. Our dashboard’s privacy-conscious design reflects this within the PHT paradigm, though choices may seem overly restrictive or lenient to others. These challenges are evident in temporal consistency assessment: in STRONG AYA, temporal data is converted to intervals (e.g. *“initial treatment occurred 30 days after diagnosis”*), blurring distinctions between atemporal and temporal plausibility. Thus, the system can only flag clearly implausible values (e.g. treatment before birth), rather than enabling sophisticated temporal analysis.

### Future directions

To address the intra-clinic aspects that the FL approach cannot detect, future work should develop a complementary dashboard, thereby establishing a uniform, scalable solution with different deployment characteristics for intra- and inter-clinic domains, applicable to other setups requiring similar quality assessment. By leveraging schema-agnostic approaches such as Flyover’s JSON-LD metadata, this would enable comprehensive quality assessment across both settings, establishing a two-step validation process in which local checks complement federated analyses.

In addition to technical integrity checks, current dashboards focus on structural dimensions (completeness, consistency, coherence) while neglecting metadata-level aspects such as documentation, provenance, coverage, and representativeness. These dimensions are becoming increasingly important under the European Health Data Space (EHDS), which requires datasets to be catalogued using standardised metadata in Health DCAT AP, Chap.IV, Article 77 (29) accompanied by standardised information on data quality and utility, Chap.IV, Article 78 (29). Incorporating such metrics would provide significant safeguards for downstream reuse while providing little extra concern in terms of privacy.

However, even with improved validation, frameworks such as Kahn et al.’s may struggle to identify statistically unusual yet valid data points (e.g. male breast cancer patients or shifts in clinical practices). Integrating machine or deep learning approaches would enable adaptive detection of population-specific anomalies, handling both structural diversity and subtle clinical patterns.

Finally, beyond the EHDS requirements themselves, initiatives such as the European Quantum label (30) are expected to operationalise standardised quality and utility reporting by providing standardised quality and utility scores that are expected to become mandatory metadata for dataset publication on national and European data portals by 2029. Schema-agnostic quality assessment frameworks will therefore be essential for regulatory compliance, dataset discoverability, and reusability. Our current approaches represent an initial step toward meeting these emerging requirements but require further refinement to align large FL projects with such efforts.

## Conclusion

Federated quality assessment systems provide essential cross-institutional oversight but may produce imperfect results for specific quality dimensions, requiring post-hoc data cleaning, while in-house approaches offer thorough validation at the cost of scalability. Through common data models like OSIRIS-RWD and OMOP, general data quality tools are indispensable for intra-clinic in-house assessment, but they often require significant refinement or data transformation to achieve true interoperability across diverse research ecosystems and only partially facilitate collaboration in FL. The optimal solution for FL ecosystems emerges from a complementary relationship between these approaches: a schema-agnostic local dashboard combined with a federated quality assessment dashboard.

For dynamic ecosystems like STRONG AYA – where data structures are diverse and continuously evolving – this integrated approach proves particularly promising. The federated system enables cross-institutional consistency checks and reveals ecosystem-wide patterns, while the in-house dashboard provides institution-specific validation and preliminary quality control. Together, they establish a robust framework that accommodates both individual dataset structures and collaborative research requirements, creating a sustainable model for data quality in evolving multi-institutional ecosystems.

## Data Availability

The data analyzed in this study is subject to the following licenses/restrictions: The datasets used are not publically available. Requests to access these datasets should be directed to Hugo Crochet.

https://github.com/STRONGAYA/federated-data-management-portal

## Acknowledgements

All individuals who have contributed to this study are included in the author list. We used generative artificial intelligence for the following tasks: Mistral AI’s Pro for typographical checking and Adobe Firefly for image generation.

## Notes

Support: J. Hogenboom, A. Sans, A.L.A.J. Dekker, W.T.A. van der Graaf, O. Husson, L.Y.L. Wee, and V. Gouthamchand are supported by the European Union’s Horizon 2020 research and innovation programme through The STRONG-AYA Initiative (Grant agreement ID: 101057482). A. Lobo Gomes is supported by Innovative Medicines Initiative (IMI), Digital Oncology Network for Europe (DigiONE), and the European Regional Development Fund (ERDF). O. Husson is also supported by the Netherlands Organization for Scientific Research through a Vidi grant (ID: 198.007). L.Y.L. Wee is also supported by ZonMW and Stichting Hanarth Fonds. N. Perez, Q. Filori, H. Crochet and A. Sans are supported by Centre Léon Bérard (Lyon, France).

### Competing Interest Statement

A.L.A.J. Dekker is employed by Medical Data Works B.V., holds stock or ownership interests in Medical Data Works B.V., and has received travel, accommodations, and expense support from Medical Data Works B.V. W.T.A. van der Graaf has receivedinstitutional research funding from Lilly.
All other authors have declared no conflicts of interest.

### Author Declarations

The study used a subset of breast cancer patients recorded in the institutional breast cancer database, restricted to patients with an information level of at least 5 - a threshold corresponding to explicit patient consent for the use of their health data in research. Secondary use of these healthcare data was conducted in accordance with the French MR004 reference methodology. The project underwent review by the data protection office of CLB (Centre Leon Berard), received a General Data Protection Regulation (GDPR) certificate, and was registered in CLB's GDPR registry. To limit re-identification risks inherent to routine clinical data reuse, we applied an internal privacy transformation involving uniform date-shifting, thereby maintaining internal temporal relationships between events. All data processing procedures are documented in the information notice available on the Unicancer transparency portal, for review by individuals included in the dataset.

