## Supplementary Table 1 for "Data Auditing and Quality Assurance in a Federated Learning Consortium; Getting the Best of Both Worlds from Cross-Institutional and In-House Data Quality Inspection"

| Dimension | Level | Subcategory | Example of quality rule | Number of existing inconsistent patients (/839) | Number of patients with inconsistencies manually added | Variables concerned | Targeted dashboard |
|---|---|---|---|---|---|---|---|
| *Consistency* | Field | Atemporal | Age at diagnosis field calculated from incidence date and birth date. Allowed values: ≥ 0 and < 100 | 0 | 5 | BirthDate/ CancerDiagnosisDate | In-house |
| *Consistency* | Field | Atemporal | Realistic Body mass Index (BMI). Allowed values: 15< BMI < 50 | 4 | 0 | Weight /Height | In-house |
| *Consistency* | Field | Atemporal | Vital status consistent with death date | 0 | 1 | DeathDate/ LastNewsStatus | In-house |
| *Consistency* | Field | Temporal | Cancer diagnosis date (date of incidence) ≥ birth date and ≤ last news date | 0 | 5 | DiagnosisDate/ LastNewsDate | In-house |
| *Consistency* | Field | Temporal | Treatment dates after cancer diagnosis date | 24 | 3 | Treatment dates | In-house |
| *Semantic* | *Concept* | - | Sex–topography code impossible combinations | 0 | 4 | TopographyCode | In-house |
| *Semantic* | *Concept* | - | Topography codes for which laterality is required but missing | 0 | 5 | Laterality / TopographyCode | In-house |
| *Semantic* | *Concept* | - | Metastatic patient at diagnosis and presence of TNM_M code at diagnosis | 0 | 1 | TNM_M / CancerDiagnosisDate | In-house |

| | | | | | | | |
|---|---|---|---|---|---|---|---|
| *Semantic* | *Concept* | - | Morphology-behaviour codes mismatch | 3 | 0 | MorphologyCode / BehaviourCode | In-house |
| *Semantic* | *Concept* | Interoperability | Primary cancer topography sites coding system e.g. ICDO-O-3 or ICD-10) | Aggregate-level only | | TopographyCode | Federated |
| *Semantic* | *Concept distribution* | Inter-organisational heterogeneity | Artificially modified laterality distribution across organisations to simulate a non-homogeneous data (non-IID) scenario | Aggregate-level only | | Laterality | Federated |

*Supplementary Table 1: Introduced inconsistencies for quality assessment*
