## Supplementary Figure 1 for "Data Auditing and Quality Assurance in a Federated Learning Consortium; Getting the Best of Both Worlds from Cross-Institutional and In-House Data Quality Inspection"

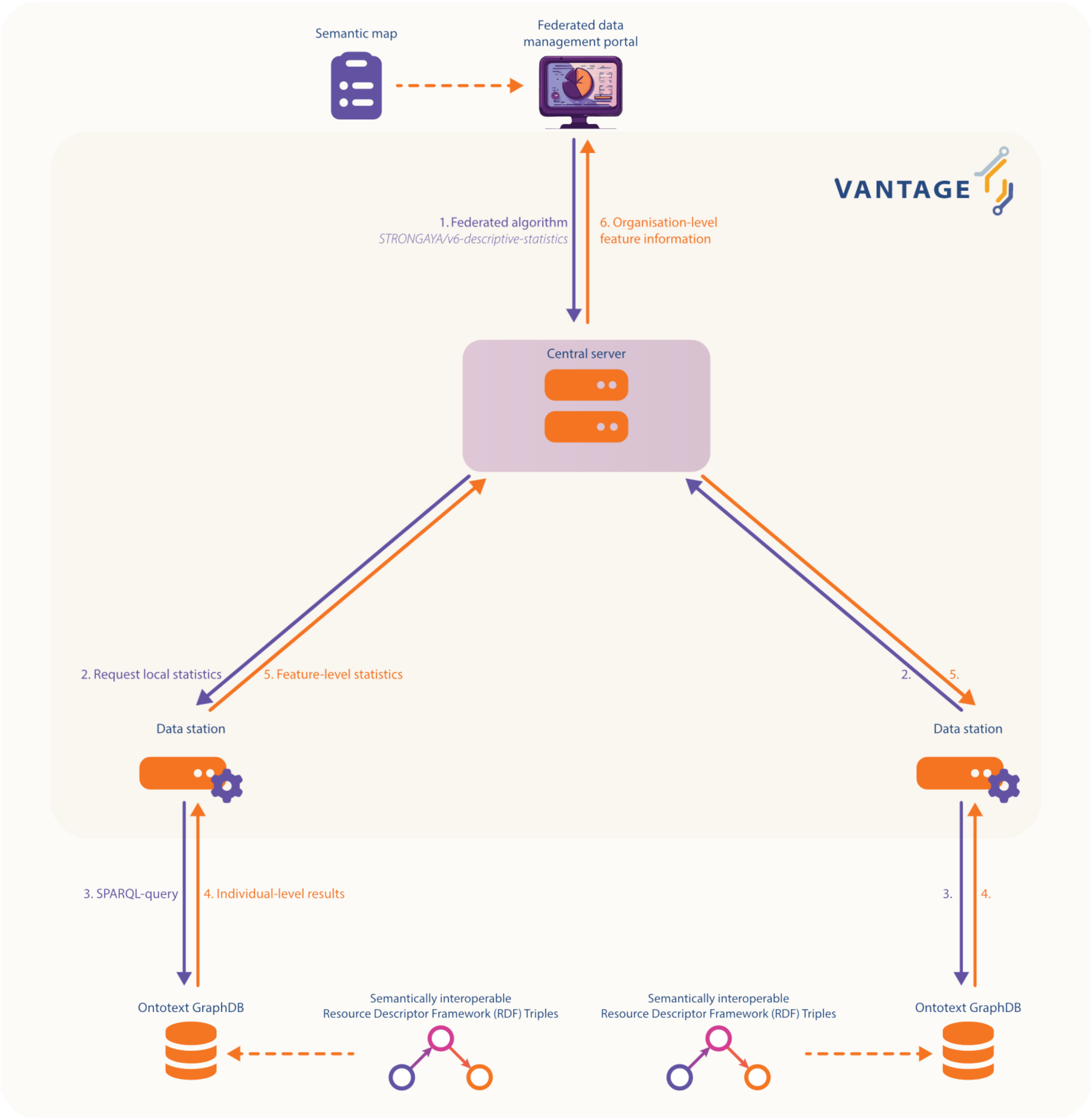


*Supplementary Figure 1: Schematic of the federated learning task integration in the dashboard and its interaction with the semantically interoperable resource descriptor framework triples.*
