## Supplementary figures and images for "Data Auditing and Quality Assurance in a Federated Learning Consortium; Getting the Best of Both Worlds from Cross-Institutional and In-House Data Quality Inspection"

### Supplementary Figure 4

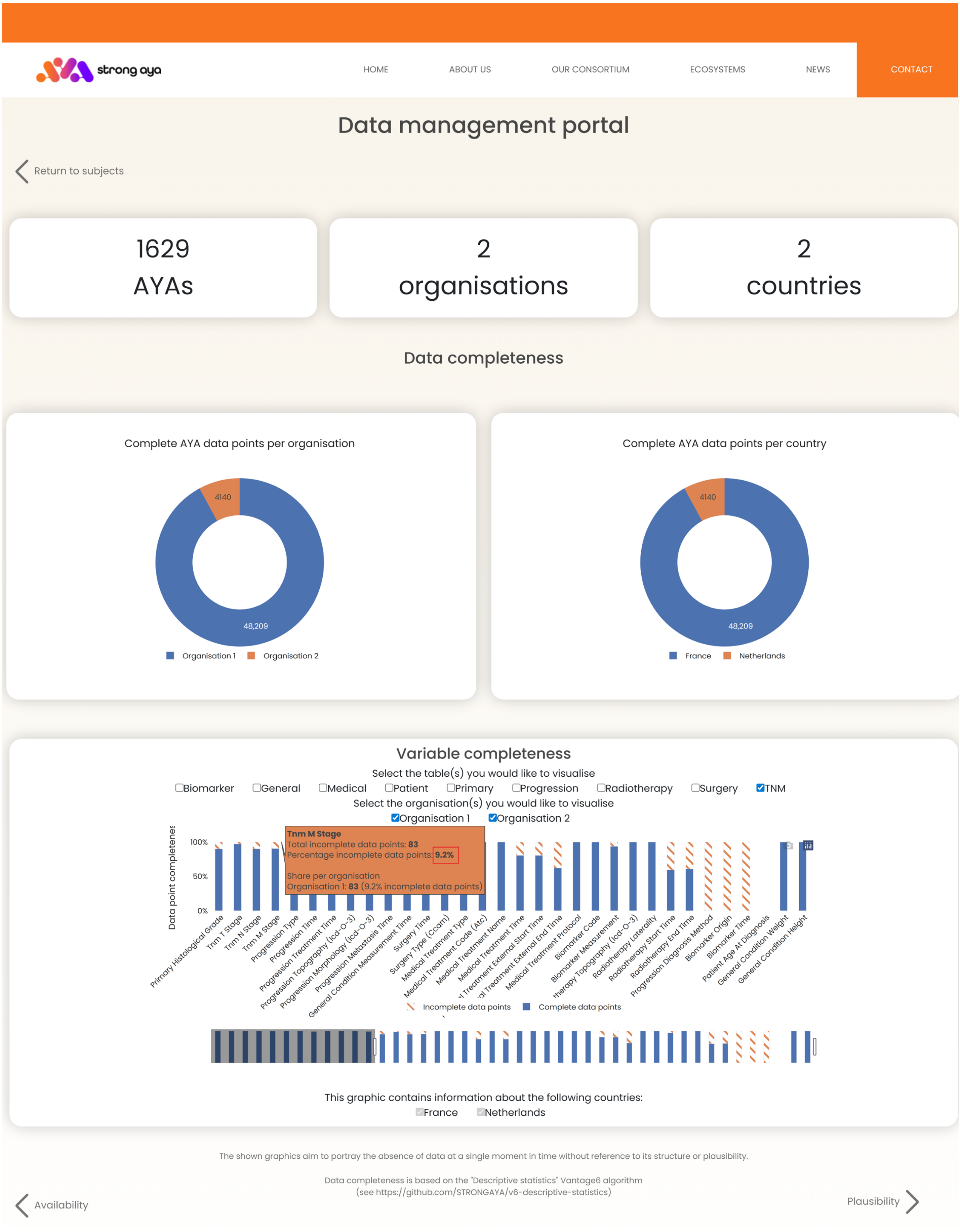


*Supplementary Figure 4: Sample of the full federated dashboard; here in data completeness view.*
