## Supplementary Figure 5 for "Data Auditing and Quality Assurance in a Federated Learning Consortium; Getting the Best of Both Worlds from Cross-Institutional and In-House Data Quality Inspection"

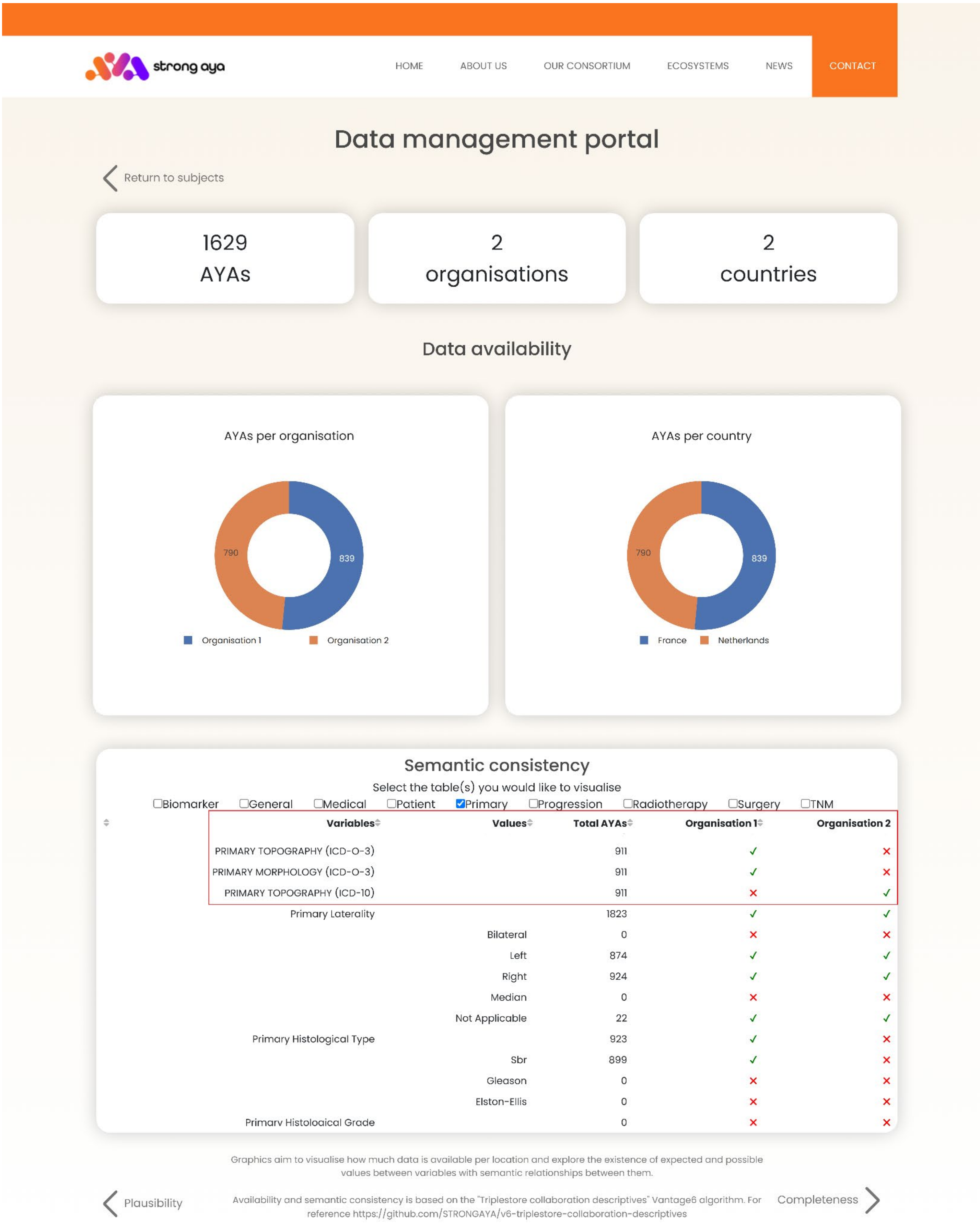


| Variables | Values | Total AYAs | Organisation 1 | Organisation 2 |
|---|---|---|---|---|
| PRIMARY TOPOGRAPHY (ICD-O-3) | | 911 | ✓ | ✗ |
| PRIMARY MORPHOLOGY (ICD-O-3) | | 911 | ✓ | ✗ |
| PRIMARY TOPOGRAPHY (ICD-10) | | 911 | ✗ | ✓ |
| Primary Laterality | | 1823 | ✓ | ✓ |
| | Bilateral | 0 | ✗ | ✗ |
| | Left | 874 | ✓ | ✓ |
| | Right | 924 | ✓ | ✓ |
| | Median | 0 | ✗ | ✗ |
| | Not Applicable | 22 | ✓ | ✓ |
| Primary Histological Type | | 923 | ✓ | ✗ |
| | Sbr | 899 | ✓ | ✗ |
| | Gleason | 0 | ✗ | ✗ |
| | Elston-Ellis | 0 | ✗ | ✗ |
| Primary Histological Grade | | 0 | ✗ | ✗ |

*Supplementary Figure 5: Sample of the full federated dashboard; here in data availability and semantic consistency view.*
